# ASSESSING THE NET FINANCIAL BENEFITS OF EMPLOYING DIGITAL ENDPOINTS IN CLINICAL TRIALS

**DOI:** 10.1101/2024.03.07.24303937

**Authors:** Joseph A. DiMasi, Abigail Dirks, Zachary Smith, Sarah Valentine, Jennifer C. Goldsack, Thomas Metcalfe, Upinder Grewal, Lada Leyens, Ute Conradi, Daniel Karlin, Lesley Maloney, Kenneth A. Getz, Bert Hartog

**Affiliations:** Tufts Center for the Study of Drug Development, Tufts University; Digital Medicine Society; F. Hoffmann-La Roche Ltd; Bayer AG; Takeda Pharmaceutical Company Limited; UCB S.A.; MindMed Inc; Genentech Inc; Janssen-Cilag B.V

**Keywords:** digital endpoints, digital health technologies, clinical trial duration, clinical trial enrollment, expected net present value, return on investment

## Abstract

**Background:** In the last few decades developers of new drugs, biologics, and devices have increasingly leveraged digital health technologies (DHTs) to assess clinical trial digital endpoints. To our knowledge, a comprehensive assessment of the financial net benefits of digital endpoints in clinical trials has not been conducted.

**Data and Methods:** We obtained data from the Digital Medicine Society (DiMe) Library of Digital Endpoints and the U.S. clinical trials registry, ClinicalTrials.gov. The benefit metrics are changes in trial phase duration and enrollment associated with the use of digital endpoints. The cost metric was obtained from an industry survey of the costs of including digital endpoints in clinical trials. We developed an expected net present value (eNPV) model of the cash flows for new drug development and commercialization to assess financial value. The value measure is the increment in eNPV that occurs when digital endpoints are employed. We also calculated a return on investment (ROI) as the ratio of the estimated increment in eNPV to the mean digital endpoint implementation cost.

**Results:** For phase 2 trials, the increase in eNPV varied from $2.2 million to $3.3 million, with ROIs between 32% to 48% per indication. The net benefits were substantially higher for phase 3 trials, with the increase in eNPV varying from $27 million to $48 million, with ROIs that were four to seven times the investment.

**Conclusions:** The use of digital endpoints in clinical trials can provide substantial extra value to sponsors developing new drugs, with high ROIs.

## INTRODUCTION

The digitization of healthcare has resulted in a wide range of new opportunities including many enhanced capabilities across the clinical trials enterprise. Digital endpoints, “a novel type of endpoint that are derived from digital health technology (DHT)-generated data (e.g. from sensors), which is often collected outside of a clinical setting, such as in a patient’s daily activities” [1] have been lauded for their promise to transform drug development. However, to date, there is a paucity of quantifiable proof points to support this claim.

In 2019, the Digital Medicine Society (DiMe) launched the open source Library of Digital Endpoints to document the use of digital endpoints to evaluate new medical products [2]. This library has grown from 38 unique digital endpoints being deployed by 12 industry sponsors [2] to 405 endpoints and 63 sponsors at the time of submission [3].

This substantial growth reflects industry recognition and embrace of the value that digital endpoints bring to clinical development. Novel digital endpoints can be both more accurate and more sensitive to meaningful functional changes and less subject to observer bias. This coupled with the ability to conduct measurements on a more or less continuous basis in a patient’s home setting make both the acquisition of the underlying data more patient friendly and more representative of real-world patient performance.

The increase in industry utilization of digital endpoints to evaluate the safety and efficacy of new medical products has been matched by the publication of evidence [4] to evaluate the performance of digital health technologies (DHTs) [5]. However, while the evidence base for high-quality measures continues to advance, there has been relatively little investigation into the financial and economic impacts of these new digital capabilities on clinical trials.

Proponents of digital health advocate for the potential of digital endpoints to reduce sample sizes and shorten study timelines [6,7]. As drug development is increasingly characterized by soaring costs [8] and trial failures driven by low accrual rates [9], the concept of de-risking against these cost drivers through the use of digital endpoints is compelling and case examples supporting these claims are emerging in the literature.

The PRESENCE study [10] leveraged digital endpoints to detect treatment effect in patients with mild to moderate Lewy Body Dementia in a phase 2 trial. The results of the PRESENCE study showed that digital measures can detect treatment effects in a smaller cohort over a shorter period than conventional clinical assessments.

With approval from the FDA, Bellerophon Therapeutics recently conducted a phase 3 study using moderate-to-vigorous physical activity (MVPA), a digital measure, as the primary endpoint. The use of MVPA helped the sponsor organization achieve a faster go/no-go decision regarding their investigational therapy [11].

Industry researchers have proposed frameworks for predicting and assessing the benefits of digital endpoints to study teams. Mori et al. developed a statistical framework to simulate the impacts of digital capabilities [12]. They applied it to trial scenarios to model potential benefits to clinical development programs. However, to our best knowledge, no field-wide assessments of the value of digital endpoints have been conducted.

Ongoing research and investment must go beyond documenting the promise of digital endpoints into documenting the value of digital endpoints. Without proving economic feasibility and financial value, the scientific advancements of these new capabilities cannot be adopted at scale.

To support the continued evaluation, development, adoption, and scale of digital endpoints, the Tufts Center for the Study of Drug Development (Tufts CSDD), an independent academic group within the Tufts University School of Medicine partnered with the Digital Medicine Society (DiMe), a global nonprofit dedicated to advancing the ethical, effective, equitable, and safe use of digital medicine to redefine healthcare and improve lives, and several industry leaders to conduct a study quantifying the net financial impact of deploying digital endpoints in clinical trials. Our goal was to evaluate whether digital endpoints are delivering on their promise and address whether digital endpoints are worth the investment.

This study presents a rigorous evaluation of the net financial benefits from including digital endpoints in clinical trials for investigational drugs in three major therapeutic areas using an expected net present value (eNPV) framework. This type of methodology has been widely applied in the pharmaceutical and other industries to assess whether investment projects are worth pursuing from a financial perspective. Through this research, we aim to drive greater field-wide visibility into the financial implications of developing and deploying digital endpoints and to support leaders within clinical development programs in decision-making as they continue to evaluate and invest in digital endpoints as a new modality in the digitization of the clinical trial enterprise.

## DATA AND METHODS

We gathered data on clinical trials for drugs, biologics, and devices that included digital endpoints among the set of outcomes that were to be evaluated according to the trial protocol from the DiMe Library of Digital Endpoints for the first quarter of 2023. Each record in the dataset represented a single digital endpoint included in the protocol for a clinical trial. The dataset contained information on 393 digital endpoints and 164 trials. Among the variables in this dataset examined for our analyses were the ClinicalTrials.gov identifying number (NCT number), if one existed, the trial study phase, trial indication, date the trial was first registered, digital endpoint, digital endpoint positioning (primary, secondary, exploratory, label claim, other), digital technology type, product type (drug, biologic, device) and trial sponsor. We placed each endpoint record in a broad therapeutic area depending on the listed indication.

We downloaded the ClinicalTrials.gov registry data as of April 14, 2023 from the Clinical Trials Transformation Initiative (CTTI) *Aggregate Analysis of Clinical Trials* (AACT) database from their website (https://aact.ctti-clinicaltrials.org/). The variables that we examined included NCT number, trial study phase, trial start date (the actual date on which the first participant was enrolled in the study), trial primary completion date (date on which the last participant in a clinical study was examined or received an intervention to collect final data for all primary outcome measures), trial enrollment, trial condition (i.e., indication), intervention type (drug, biologic, device, and other types), intervention name, sponsor class (industry, NIH, other federal agency, other governments, network, other sponsor types), lead or collaborator (whether the sponsor class value is for the lead sponsor or is a collaborator, sponsor name, study type (interventional, observational, observational with patient registry, and expanded access), number of countries involved (overall and by region), and the number of trial sites (overall and by region). Data from the DiMe Library of Endpoints and the ClinicalTrials.gov datasets were merged for analysis by the NCT identifier.

### Inclusion criteria

The ClinicalTrials.gov dataset that we downloaded contains 448,445 trial records (Figure 1). We restricted the number for analysis according to five inclusion criteria. The earliest listing for a digital endpoint in the endpoints dataset is 2005. So, we limited the trial registry records to those for which the trials were initiated in 2005 or later (n=414,264). We also restricted the analysis to interventional studies (n=345,907). We further restricted the analysis to drugs, biologics, and device trials (n=232,662). The DiMe Library of Endpoints dataset is industry-based. So, we restricted the analysis to those records in the trial registry for which industry was the lead sponsor and/or a collaborator (n=136,602). Finally, the endpoints dataset is concentrated in three therapeutic areas (Supplemental Figure S1). Central nervous system trials (CNS) accounted for 35.4% of the records in the endpoints dataset, while diabetes trials accounted for 20.7% and cardiovascular trials accounted for 14.0%, for a total of 70.1% of the dataset. Other categories had much smaller shares. Restricting the registry data to trials with indications that matched or closely matched the indications in the endpoints dataset in these three areas resulted in 48,765 registry trials.

**Figure 1.**
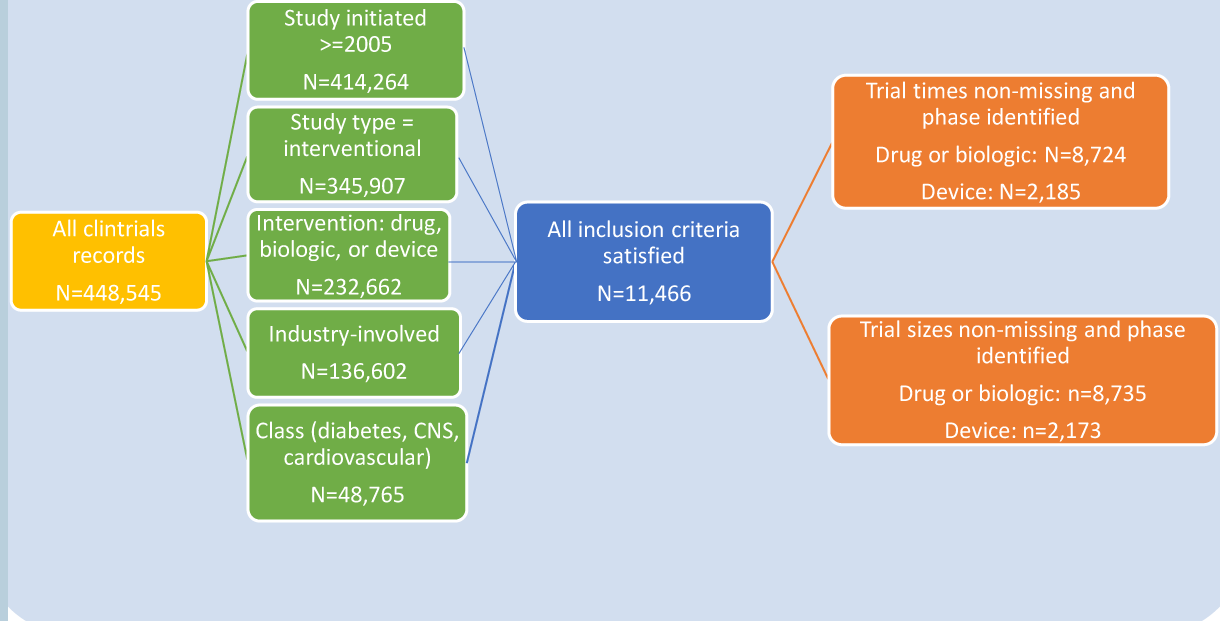
Study Inclusion Criteria and Sample Size

Our analyses were conducted on a restricted dataset that met all five inclusion criteria (n=11,466). The dataset was further reduced marginally when records with missing data for our operational metrics were excluded (Figure 1). Devices accounted for less than one-quarter of the final analysis dataset. Distinguishing between drugs and biologics is problematic because there is no easy way to identify the intervention of interest in the registry trials and because of reduced sample sizes for the trials with digital endpoints. So, we combined drugs and biologics for analysis and, for ease of reference, we henceforth refer to all those cases as drug trials. Descriptive and inferential statistical analyses were conducted by using SAS^®^ 9.4 software.

### Benefits

The potential benefits to utilizing digital endpoints in clinical trials were measured by differences in average clinical trial durations and trial enrollment sizes between studies that leveraged digital endpoints and studies that did not. Trial duration was defined as time from study start to primary completion. As noted, these dates are taken from the clinical trials registry data.

We provide comparative descriptive statistics for trials with and without digital endpoints by clinical trial phase and the three therapeutic areas noted above.^1^ While this is instructive, the differences between the two groups could be affected by a number of confounding factors. Thus, for drug trials we also analyzed the data by specifying clinical phase least squares regressions to be estimated, where the dependent variable (trial cycle time or trial size) was a function (ordinary or semi-logarithmic least squares regressions) of a benefit metric and the independent (explanatory) variables included dummy variable values for categorical variables for the presence of digital endpoints and by therapeutic class, and continuous variables for the number of trial sites, the number of trial endpoints, and a yearly trend variable.^2^

To optimize model selection by clinical phase we chose final model specifications for the financial analysis by running regressions using three model selection techniques. Specifically, we used the backward elimination, forward selection, and stepwise selection techniques in SAS^®^ 9.4. We selected model variables for inclusion in the benefit analysis if the variable was among the final set of factors (i.e., the set of variables for which the model as a whole had the most statistically significant predictive power) for at least two of the three model selection techniques.

The data were deemed sufficiently large for phase 2 and phase 3 drug trials. We do not have financial modeling data for phase 4 or device studies. Because of uncertainties in projections at early development stages, the financial valuation technique (eNPV) is typically not applied earlier than phase 2. Thus, the application of our financial analysis is restricted to phase 2 and phase 3 drug development. All parameter costs and returns were converted to constant 2023 dollars using the GDP Implicit Price Deflator as the price index.

### Implementation costs

We expect that any financial benefits from improved operational characteristics may be at least partially offset by the costs of developing, validating, and implementing DHTs to gather information on digital endpoints in clinical trials. We were not aware of any comprehensive published data on such costs. To remedy this and complete our modeling, Tufts CSDD and DiMe conducted a survey of clinical trial sponsors and developers of digital measures and digital measurement products to gather information on costs.

Responses were collected from two separate versions of the survey, one tailored to trial sponsors and the other to developers of digital measures and measurement products that are used to gather data for digital measures. The survey was administered from July 2023 to August 2023. We obtained 38 responses from the sponsor survey and 42 responses from the developer survey. Information was requested on product type, therapeutic area, endpoint type, DHT type, reasons for including digital endpoints, involvement in development, whether the measures were analytically and/or clinically validated, and costs incurred. Cost data were limited, with seven responses for technology developer costs and 11 responses for trial sponsor costs. Means, medians, and ranges were calculated for costs. To be consistent with eNPV methodology and because the focus here is on sponsor financial benefit, mean sponsor cost was used for the base case financial analyses, but we also examined what the results would be if median cost were applied to the model.

### Expected Net Present Value Framework and ROI

We utilized a version of a widely accepted methodology for quantifying the value of an investment project and determining if the investment is worth pursuing in a purely financial sense. Our eNPV framework is a lifecycle model of pharmaceutical industry drug development and commercialization. It is a risk-adjusted discounted cash flow analysis discounted to either the start of phase 2 or the start of phase 3 testing.

The model accounts for the risk of failure in development by therapeutic area. It includes estimates and assumptions about both pre- and post-approval R&D costs, the number of pre-approval indications investigated, and the number of post-approval approved supplemental indications by therapeutic area.

Other parameters needed for the model are estimates and assumptions about the cost of capital, peak sales, years to peak sales, the shape of a net revenue curve, an exclusivity period, market share erosion after generic entry, launch costs, marketing and sales costs, costs of goods sold, medical affairs, other operating costs, and taxes.

Some of the authors have used the basic eNPV framework for a number of pharmaceutical applications. These include the net benefits of patient engagement methods in clinical trials, single-source manufacturing, incremental formulation and real-time manufacturing during clinical trials, and the use of decentralized clinical trials [13–16]. The reader can find further details about the basic methodology used here in these earlier studies.

The perspective of the analysis can be done at different levels. Some of these prior studies were done on a per approved molecule basis [14–16]. That is, the net benefits are measured as if the purported improvement in drug development is applied to all development for a molecule and valued on a per molecule basis. Here, as in [13], the perspective is on a per approved indication basis. Thus, development costs, risks, and returns are determined at the indication level.

The value of including digital endpoints in clinical trial protocols was measured here as the increment in eNPV when digital endpoints are employed, as opposed to when no digital endpoints are applied for the same or similar indication. We developed a return on investment metric (ROI) for utilizing digital endpoints in clinical trials as the ratio of the increment in eNPV from using digital endpoints to the mean implementation cost of including digital endpoints in clinical trials, as reported by clinical trial sponsors.

## RESULTS

The DiMe Library of Endpoints is a rich source of information on digital endpoints in industry clinical trials. Using this dataset we can characterize some of the major attributes of digital endpoints to date. Our dataset, for example, characterizes the positioning of the endpoints (Supplemental Figure S8). Nearly one-third (31.8%) of the digital endpoints were primary outcomes, while somewhat more than half (56.0%) were secondary outcomes. The data also include information on technology types associated with the digital endpoints (Supplemental Figure S9). More than half (55.0%) of the DHTs are connected sensors. Nearly half (49.4%) of the trials with digital endpoints had more than one digital endpoint (Supplemental Figure S10). 11.5% of the trials had six or more digital endpoints.

### Quantifying Digital Endpoint Benefits

Table 1 presents phase duration means and medians for trials that leverage digital endpoints along with thos that do not. This table presents data acoss the three therapeutic areas included in the analysis by clinical phase for drugs and devices. Although the results are somewhat mixed, in the vast majority of cases trials with digital endpoints tended to have shorter trial durations. For phase 2 trials both the means and medians for trial cycle time are lower for trials with digital endpoints compared to trials with no digital endpoints for CNS and diabetes drugs. Trial durations were 7.9 and 6.0 months lower for CNS trials with digital endpoints. For diabetes trials the means and medians were 6.0 and 6.4 months shorter, respectively. For cardiovascular trials the mean and median trial durations were somewhat higher for the digital endpoint trials.

**Table 1.**
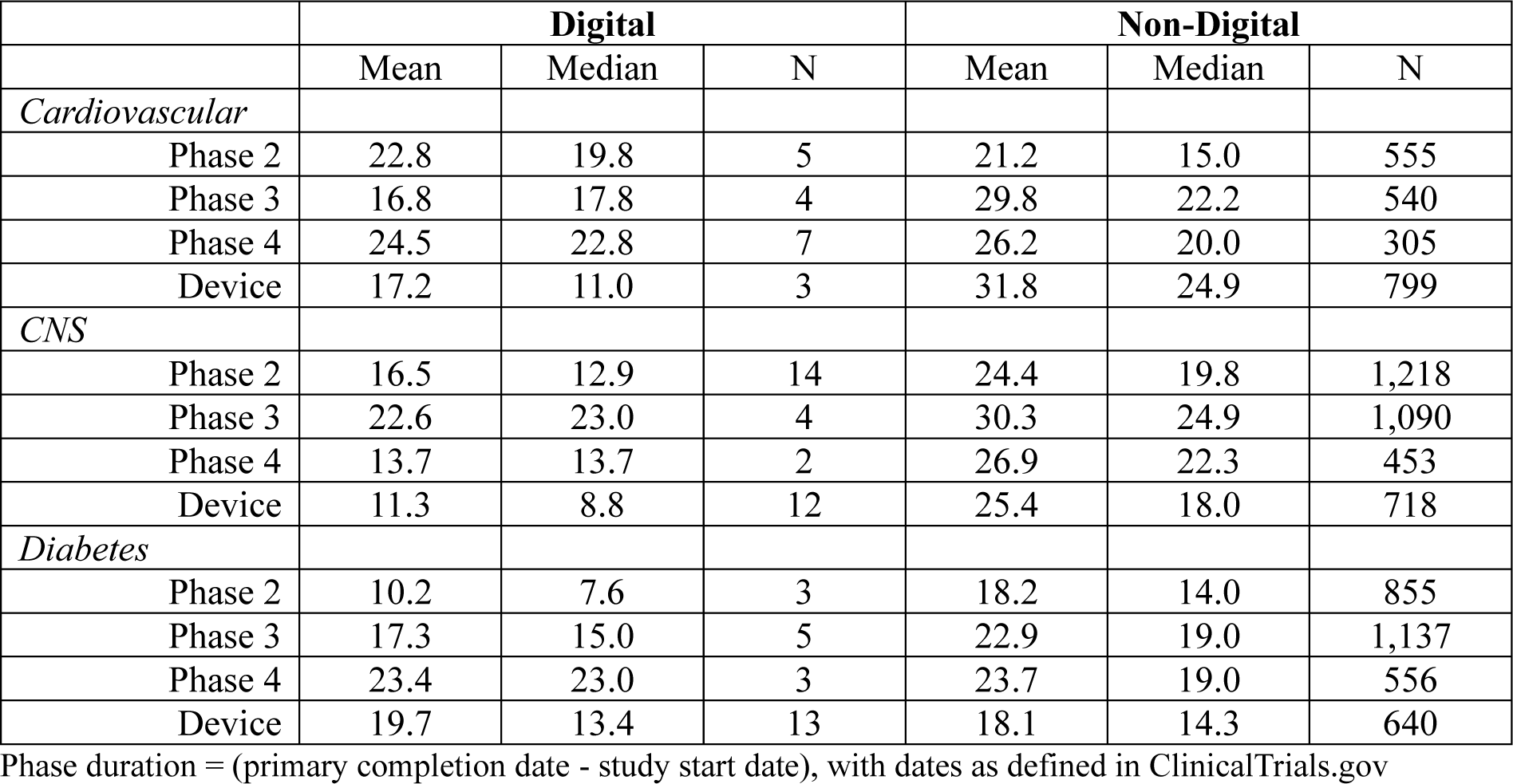
Phase duration for trials with and without digital endpoints by therapeutic area.

|  | **Digital** |  |  | **Non-Digital** |  |  |
| --- | --- | --- | --- | --- | --- | --- |
|  | Mean | Median | N | Mean | Median | N |
| *Cardiovascular* |  |  |  |  |  |  |
| Phase 2 | 22.8 | 19.8 | 5 | 21.2 | 15.0 | 555 |
| Phase 3 | 16.8 | 17.8 | 4 | 29.8 | 22.2 | 540 |
| Phase 4 | 24.5 | 22.8 | 7 | 26.2 | 20.0 | 305 |
| Device | 17.2 | 11.0 | 3 | 31.8 | 24.9 | 799 |
| *CNS* |  |  |  |  |  |  |
| Phase 2 | 16.5 | 12.9 | 14 | 24.4 | 19.8 | 1,218 |
| Phase 3 | 22.6 | 23.0 | 4 | 30.3 | 24.9 | 1,090 |
| Phase 4 | 13.7 | 13.7 | 2 | 26.9 | 22.3 | 453 |
| Device | 11.3 | 8.8 | 12 | 25.4 | 18.0 | 718 |
| *Diabetes* |  |  |  |  |  |  |
| Phase 2 | 10.2 | 7.6 | 3 | 18.2 | 14.0 | 855 |
| Phase 3 | 17.3 | 15.0 | 5 | 22.9 | 19.0 | 1,137 |
| Phase 4 | 23.4 | 23.0 | 3 | 23.7 | 19.0 | 556 |
| Device | 19.7 | 13.4 | 13 | 18.1 | 14.3 | 640 |

Phase duration = (primary completion date - study start date), with dates as defined in ClinicalTrials.gov

For phase 3 trials the trial duration averages were uniformly shorter for trials with digital endpoints, ranging from a low of a 1.9 month decrease for median CNS trials to a high of a 13.0 month lower mean for cardiovascular trials. Phase 4 trial duration means were lower for digital endpoint trials for all three therapeutic areas, ranging from 0.3 months lower for diabetes trials to 13.2 months for CNS trials. The median durations were lower (8.6 months) only for CNS trials with digital endpoints. For device trials both the mean and median trial durations were lower for trials with digital endpoints for cardiovascular and CNS indications. However, there was little difference in mean and median durations for diabetes device trials that leveraged digital endpoints compared to those that did not.

Table 2 contains our results for trial enrollment sizes by phase and therapeutic area for trials with and without digital endpoints. Here, the results also show, for the most part, smaller trial sizes when digital endpoints are employed. For phase 2 trials mean and median enrollment sizes were lower for CNS and diabetes trials (87.4 and 73.0 subjects on average, respectively) that leveraged digital endpoints.

**Table 2.** Phase enrollment for trials with and without digital endpoints by therapeutic area.

|  | **Digital** |  |  | **Non-Digital** |  |  |
| --- | --- | --- | --- | --- | --- | --- |
|  | Mean | Median | N | Mean | Median | N |
| *Cardiovascular* |  |  |  |  |  |  |
| Phase 2 | 151.5 | 116.5 | 4 | 123.3 | 63.0 | 547 |
| Phase 3 | 354.0 | 394.5 | 4 | 809.6 | 302.0 | 526 |
| Phase 4 | 95.1 | 66.0 | 7 | 253.0 | 84.0 | 297 |
| Device | 175.0 | 205.0 | 3 | 776.3 | 62.0 | 782 |
| *CNS* |  |  |  |  |  |  |
| Phase 2 | 51.5 | 41.0 | 13 | 138.9 | 78.0 | 1,206 |
| Phase 3 | 286.8 | 237.0 | 4 | 571.8 | 343.0 | 1,118 |
| Phase 4 | 58.0 | 58.0 | 2 | 237.2 | 93.0 | 445 |
| Device | 36.9 | 40.0 | 12 | 113.3 | 39.0 | 707 |
| *Diabetes* |  |  |  |  |  |  |
| Phase 2 | 80.3 | 42.0 | 3 | 153.3 | 96.0 | 851 |
| Phase 3 | 381.2 | 393.0 | 5 | 646.1 | 376.5 | 1,141 |
| Phase 4 | 61.3 | 59.0 | 3 | 369.5 | 102.0 | 555 |
| Device | 82.4 | 80.0 | 13 | 128.2 | 60.0 | 656 |

Enrollment sizes taken from ClinicalTrials.gov trial records.

For phase 3 trials mean trial size was lower across all three therapeutic areas for trials that leveraged digital endpoints, and the reduction in trial size associated with the use of digital endpoints was greater in absolute terms for phase 3 trials compared to phase 2 trials. The mean reductions in phase 3 trial sizes for trials with digital endpoints was 455.6, 285.0, and 264.9 for cardiovascular, CNS, and diabetes trials, respectively. The results for medians were mixed.

Phase 4 mean and median trial sizes were all lower for trials with digital endpoints. Mean reductions in phase 4 trial sizes for trials with digital endpoints were 157.9, 179.2, and 308.2 for cardiovascular, CNS, and diabetes trials, respectively. For device trials mean, but not median, trial sizes were lower for trials with digital endpoints. Mean trial sizes were lower by 601.3, 76.4, and 45.8 subjects for cardiovascular, CNS, and diabetes trials, respectively.

These descriptive statistics are informative, but they may be affected by differences in confounding factors. For example, we found that digital endpoint trials tend to have used more sites per trial than did non-digital trials, particularly when extreme outliers are excluded (Supplemental Figures S11-S12). Additionally, we found that, on average, digital endpoint trials had protocols with more endpoints than did non-digital trials (Supplemental Figures S13-S14). Finally, we noted trends in trial durations and enrollments for trials as a whole, and the distribution of trials by year differs somewhat between trials with and without digital endpoints. So, we examined these factors in the context of regression models for trial duration and trial size, along with variables that distinguish by therapeutic area and whether a trial included digital endpoints or not.

We selected model variables for inclusion in ordinary least squares and semi-log regressions for trial duration and trial size by applying backward elimination, forward selection, and stepwise selection techniques. The variables that were included in the final runs for at least two of these techniques were used to estimate duration and trial size advantages associated with the inclusion of digital endpoints in clinical trials.

The results indicated that the final set of regression independent variables include categorical variables for whether a trial had digital endpoints, the trial indication’s therapeutic area, the year in which the trial started, the number of trial sites, and the number of trial endpoints. The analysis showed that the best specifications were semi-logarithmic for phase 2 and phase 3 duration and phase 3 enrollment, while ordinary least squares was best for phase 2 enrollment.

The coefficients for the independent variables for the various duration and enrollment regressions are given in Supplemental Table S1. Although the results would be statistically significant if the number of sites were included in the enrollment regressions, we reasoned that the number of sites should be correlated with trial sizes but that trial size is not determined by the number of sites. Instead, trial size often informs the number of sites. So, we did not include the number of sites as potential explanatory variables in the enrollment regressions.

The estimated coefficients and the regression model specification can be applied to determine estimates of the reduction in trial duration and trial size by therapeutic area and phase when digital endpoints are employed. The results, when the predicted values for the dependent variables are estimated at mean values for the continuous explanatory variables (Supplemental Table S2), are shown in Table 3. With semi-logarithmic specifications the percentage decline in durations and sizes are restricted to be constant across therapeutic areas, while the absolute declines are allowed to be variable. For ordinary least squares regressions, as here for phase 2 enrollment, the absolute declines will be constant, while percentage declines can be variable.

**Table 3.** Predicted reductions in trial duration (mos.) and size with digital endpoints by therapeutic area.

|  | Phase 2 Duration |  |  | Phase 3 Duration |  |  | Phase 2 Enrollment |  |  | Phase 3 Enrollment |  |  |
| --- | --- | --- | --- | --- | --- | --- | --- | --- | --- | --- | --- | --- |
|  | Predicted value | Absolute decline | Percent decline | Predicted value | Absolute decline | Percent decline | Predicted value | Absolute decline | Percent decline | Predicted value | Absolute decline | Percent decline |
| Cardiovascular |  |  |  |  |  |  |  |  |  |  |  |  |
| *Non-digital* | 14.8 |  |  | 21.9 |  |  | 93.0 |  |  | 181.2 |  |  |
| *Digital* | 11.2 | 3.5 | 24.0% | 17.0 | 4.9 | 22.0% | 77.7 | 15.2 | 16.4% | 160.0 | 21.2 | 11.7% |
| CNS |  |  |  |  |  |  |  |  |  |  |  |  |
| *Non-digital* | 17.4 |  |  | 23.9 |  |  | 109.3 |  |  | 277.0 |  |  |
| *Digital* | 13.3 | 4.2 | 24.0% | 18.7 | 5.2 | 22.0% | 94.1 | 15.2 | 13.9% | 244.6 | 32.4 | 11.7% |
| Diabetes |  |  |  |  |  |  |  |  |  |  |  |  |
| *Non-digital* | 13.7 |  |  | 18.2 |  |  | 130.8 |  |  | 312.6 |  |  |
| *Digital* | 10.4 | 3.3 | 24.0% | 14.2 | 4.0 | 22.0% | 115.6 | 15.2 | 11.6% | 276.0 | 35.6 | 11.7% |

Predicted values for dependent variables determined at mean values for the continuous explanatory variables
Phase duration = (primary completion date - study start date), with dates as defined in ClinicalTrials.gov

The results suggest that the use of digital endpoints in clinical trials were associated with average declines in phase 2 duration of 3.5, 4.2, and 3.3 months for cardiovascular, CNS, and diabetes trials, respectively. Similarly, average phase 3 durations were estimated to be 4.9, 5.2, and 4.0 months lower for trials with digital endpoints across cardiovascular, CNS, and diabetes trials, respectively. We apply these results to an eNPV model that is parameterized by phase-to-phase times (i.e., the start of one phase to the start of the next phase). We made the assumption that the phase-to-phase times will be reduced by the same amount as the reductions in trial times that we found. Since the basic measurement unit in our eNPV model is a month, we apply the duration time reduction results to our model by rounding them to the nearest month. So, in applying the eNPV model we assume for phase 2 that phase-to-phase times are reduced by 4,4, and 3 months for cardiovascular, CNS, and diabetes phase-to-phase times, respectively.

Similarly, for phase 3 we assume that phase-to-phase times are reduced by 5, 5, and 4 months for cardiovascular, CNS, and diabetes phase-to-phase times, respectively.

We do not have trial enrollment sizes among the parameterized values in our eNPV model, but we do have clinical phase costs. We assume that phase costs are proportional to trial sizes. So, we use the results on the estimated percentage declines in trial sizes for phase 2 and phase 3. Thus, the benefit of lower trial sizes when digital endpoints are used are manifested in our model through lower development costs. So, we assume that trial sizes and phase costs decline by 16.4%, 13.9%, and 11.6% when digital endpoints are used for cardiovascular, CNS, and diabetes trials, respectively. For phase 3, we assume that enrollment and phase cost declines by 11.7% for each therapeutic area when digital endpoints are included in trials.

### Digital Endpoint Costs

We surveyed trial sponsors and developers of digital measures and digital measurement products to gather information on the development, validation, and utilization of digital endpoints in clinical trials. Costs were reported by year, so that we can convert the cost data to constant dollars. The data were limited, but as far as we know, this is the first evidence presented publicly on the costs of the development and use in clinical trials of these new measures.

From the survey shared with developers of digital measures and digital measurement products, we found, for nine cases, that 77.8% of the digital measures were both analytically and clinically validated, 11.1% were analytically but not clinically validated, and 11.1% were neither analytically nor clinically validated. Total costs incurred by developers associated with the development of digital measures (n=7) ranged (in constant 2023 dollars) from $348,429 to $120,988,950. The mean total cost was $21,042,334, while the median cost was $3,791,378. Developers also reported the value of agreements with trial sponsors for the use of their measures and/or measurement products to collect digital endpoints. Across 10 developer responses, the mean value of these agreements was $783,258 while the median value of these agreements was $586,039.

From the survey designed for sponsors we found, for 12 sponsor survey cases, that the digital health measures leveraged as endpoints in their studies were both analytically and clinically validated for 41.7% of the cases, analytically but not clinically validated for another 41.7% of the cases, and neither analytically nor clinically validated for 16.7% of the cases. With respect to our eNPV model, though, we were most interested in the additional costs incurred by sponsors associated with the implementation of digital endpoints in their studies. These results are shown in Supplemental Figure S15. The mean cost was $3,416,060 and the median cost was $1,000,000. We use the mean cost for our base value analyses as that is consistent with the eNPV model and it provides a conservative measure of value, but in sensitivity analyses we examine what the results would be using the median cost.

### Base Case Parameterizations for Digital Endpoint eNPV Models

We applied the results found for this study on the benefits and costs of utilizing digital endpoint measures to our eNPV model. Most of the industry parameters modeled, and values for them, are shown in a previous study of the value of decentralized clinical trials [16]. Sources for key parameters are shown in Supplemental Table S3. However, critical differences between this study and the previous study are that we take the perspective here of value at the indication level, as opposed to the molecule level, and we need to differentiate among therapeutic areas. In particular, we need here to choose values for the number of indications pursued pre-approval and the number of indications approved post-approval, and values to account for differences in development costs and technical development risks by therapeutic area.

We assume that two indications are investigated prior to original approval for each therapeutic area. For purposes of calculating implementation costs, we assume two phase 2 trials and two phase 3 trials per pre-approval indication. These assumptions can easily be varied in sensitivity analyses. For indications approved after original approval, we examine approvals data at the Food and Drug Administration (FDA) website. Supplemental Figure S16 shows the average number of indications approved by the FDA for drugs originally approved by the FDA during 2007-2018. Results are shown there for 2007-2014 and the entire period, 2007-2018. The more recent original approvals have not had as much time for supplemental indication approvals to occur. So, it is more appropriate to use the early period, 2007-2014. Therefore, for our eNPV model we assume, on average, two indication approvals for cardiovascular drugs, 2.22 indication approvals for CNS drugs, and 2.78 approvals for diabetes drugs. We need these assumptions for the revenue analysis. We collected annual sales data from the Cortellis™ pipeline database, which contains information on consensus analyst forecasts. But these are total sales data for the molecule. To get annual sales per indication we divide by the average number of indications for drugs in the three therapeutic areas. These calculations yielded per indication peak sales of $836,275,00 for cardiovascular indications, $482,459,000 for CNS indications, and $1,007,006,000 for diabetes indications.

We take phase transition and clinical approval success rates at the indication level for our three therapeutic areas from a recent study [17]. The results are shown in Supplemental Table S4. Finally, we need to differentiate clinical phase costs and phase-to-phase durations by therapeutic area for a base case. Trial budget information from the Tufts CSDD Protocol Complexity Benchmark Database and cost and time results from an industry R&D cost study [18] were used to find relative phase costs and durations for the three therapeutic areas. The results are shown in Supplemental Table S5. This allows us to vary cost and time parameters in the base cases for the three therapeutic areas.

### Digital Endpoint Value for Phase 2 Trials

We applied the estimated benefits and costs to our eNPV model and determined the change in eNPV when digital endpoints are employed in phase 2 clinical trials compared to the base case eNPVs for the three therapeutic areas on which we focus. A positive change in eNPV (eNPV delta) indicates that the proposed alternative to the base case is worth pursuing from a purely financial perspective.

The results are shown in Table 4. Given the results above, we assumed that the introduction of digital endpoints in clinical trials results in reductions in time from phase 2 start to phase 3 start of four months for cardiovascular and CNS indications and three months for diabetes indications. Additionally, we assumed that trial sizes were lower for phase 2 trials with digital endpoints by 11.0% for cardiovascular trials, 13.9% for CNS trials, and 16.4% for diabetes trials. For all therapeutic areas, there is a gain in eNPV at the mean cost of implementing digital endpoints. The eNPV deltas were $2.2 million for cardiovascular indications, $2.1 million for CNS indications, and $3.3 million for diabetes indications. The ROIs for implementing digital endpoints in phase 2 trials were 32.4% for cardiovascular indications, 30.5% for CNS indications, and 47.7% for diabetes indications.

**Table 4.** Base case change in eNPV (thousands 2023 USD) and ROI per phase 2 investigational indication for digital endpoint clinical trials by therapeutic area.

|  |  |  | Mean implementation cost |  | Median implementation cost |  |
| --- | --- | --- | --- | --- | --- | --- |
| *Therapeutic area* | *Reduction in phase duration (mos.)* | *Reduction in trial size* | *eNPV delta* | *ROI* | *eNPV delta* | *ROI* |
| Cardiovascular | 4 | 11.0% | \$2,214 | 0.324x | \$5,983 | 3.0x |
| CNS | 4 | 13.9% | \$2,086 | 0.305x | \$5,797 | 2.9x |
| Diabetes | 3 | 16.4% | \$3,256 | 0.477x | \$6,967 | 3.5x |

Mean implementation cost (thousands 2023 USD) per trial = \$3,416; median implementation cost (thousands 2023 USD) per trial = \$1,000
Costs and returns discounted to the start of phase 2 testing
ROI = eNPV/sponsor implementation cost per indication (assumes two phase 2 trials per indication)

Table 4 also shows what the results would be if we used the median implementation cost reported by sponsors ($1.0 million). The results are substantially better. This happens because the median cost is lower than the mean cost. As a result, the eNPV delta will be higher because of lower resource costs and, additionally, the implementation cost is the denominator of the ROI metric. The eNPV deltas are 2.1 to 2.8 times higher than when the mean cost is used. The ROI values are 2.2 to 2.4 times higher.

### Digital Endpoint Value for Phase 3 Trials

We also examined results under the assumption that digital endpoints are applied only in phase 3 trials. The results are shown in Table 5. Here we assumed that the introduction of digital endpoints in clinical trials results in reductions in time from phase 3 start to regulatory submission of five months for cardiovascular and CNS indications and four months for diabetes indications. Additionally, we assumed that trial sizes were 11.7% smaller for phase 3 trials with digital endpoints across all three therapeutic areas.

**Table 5.** Base case change in eNPV (thousands 2023 USD) and ROI per phase 3 investigational indication for digital endpoint clinical trials by therapeutic area.

|  |  |  | Mean<br>implementation cost |  | Median<br>implementation cost |  |
| --- | --- | --- | --- | --- | --- | --- |
| *Therapeutic area* | *Reduction in phase duration (mos.)* | *Reduction in trial size* | *eNPV delta* | *ROI* | *eNPV delta* | *ROI* |
| Cardiovascular | 5 | 11.7% | \$33,280 | 4.9x | \$36,836 | 18.4x |
| CNS | 5 | 11.7% | \$27,344 | 4.0x | \$30,971 | 15.5x |
| Diabetes | 4 | 11.7% | \$48,404 | 7.1x | \$52,105 | 26.1x |

Mean implementation cost (thousands 2023 USD) per trial = \$3,416; median implementation cost (thousands 2023 USD) per trial = \$1,000
Costs and returns discounted to the start of phase 3 testing
ROI = eNPV/sponsor implementation cost per indication (assumes two phase 3 trials per indication)

The increments in eNPV at the mean implementation cost are much higher than they are for phase 2, ranging from $27.3 million to $48.4 million. One reason why the measured increase in value is higher is that the probability that an investigational drug that has entered phase 3 will be approved is much higher than for a phase 2 investigational drug (Supplemental Table S4).

Consequently, the benefits of shorter development times from earning revenues sooner are greater for phase 3 because expected revenues (probability of approval multiplied by revenues if a drug is approved) are higher. The ROIs for phase 3 were also much higher, ranging from 4.0 times the total investment (implementation) cost to 7.1 times the investment cost.

If median implementation costs are used, the eNPV deltas are, as expected, higher, ranging from 8% to 13% higher. The ROIs are 3.7 to 3.9 times higher when median, as opposed to mean, implementation cost is used. The results in aggregate demonstrate that incorporating digital endpoints in clinical trials provides substantial extra financial value to sponsors across all therapeutic areas considered and for both phase 2 and phase 3.

### Sensitivity Analysis

Any of the numerous model parameter values can be changed and results analyzed. Given the assumed benefits in trial durations and trial sizes, unless the changes in parameter values are extreme, they generally support the conclusion that incorporating digital endpoints in clinical trials yields substantial financial value. Here, we focus on sensitivity analyses for the digital endpoint benefit values. That is, we suppose that reductions in trial durations and trial sizes are lower or higher than our base case estimated values.

The panels in Figure 2 show ROI at varying assumed reductions in trial durations holding base case reductions in trial size constant for phase 2 and phase 3 trials and by therapeutic area (Figure 2a and Figure 2c). We also examined the sensitivity of ROI for varying assumptions about percentage reductions in trial sizes, holding base case reductions in phase duration constant for phase 2 and phase 3 trials and by therapeutic area (Figure 2b and Figure 2d). In this way, we can isolate the effects on financial value for the two types of benefit.

**Figure 2.**
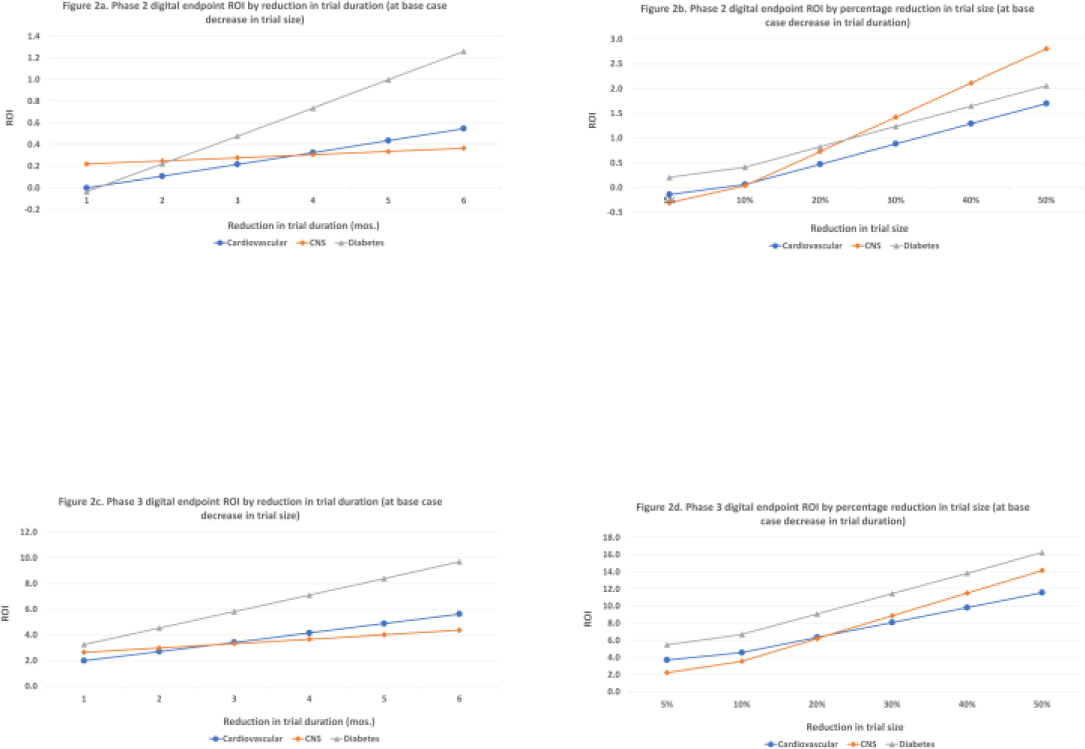
Sensitivity Analyses for Phase Duration and Trial Size by Phase and Therapeutic Area (at mean implementation cost)

The results show generally higher ROIs for diabetes indications compared to cardiovascular and CNS indications. The exceptions are phase 2 reductions in trial sizes, holding phase duration constant, for high assumed percentage reductions in trial sizes (30% or more in relation to CNS trials). ROIs for phase 3 trials are at least twice the investment cost for both trial duration variation and trial size variation. For phase 2, with the exception of CNS trials, at very low reductions in phase duration or trial size (one month for phase duration reduction and 5% for trial size reduction) the ROIs become zero or negative. Finally, the ROIs are much higher in all cases, and uniformly positive, if median implementation cost is used in the modeling (Supplemental Figure S17).

## DISCUSSION

Our research shows that the development of digital endpoints, and the DHTs that underlie them, require substantial upfront investment. However, these data also show that there is a significant return on that investment. Stakeholders across the clinical trials enterprise have the opportunity to capture even greater value through the scale of these capabilities and increase overall returns through reuse and repeatability. A cross-portfolio digital measures strategy that encompasses both digital biomarkers and digital endpoints should be a core component of research and development strategies across therapeutic areas to optimize this increased value.

As this analysis suggests, the returns associated with deploying digital measures in clinical trials are substantive. One opportunity to further dilute costs and increase overall returns for organizations investing in these capabilities is for those organizations to engage in precompetitive collaborations. These forums not only enable organizations to share the risk of digital measure development but result in standardized approaches to measure development and harmonization of new measures across clinical research and/or clinical care. Such forums also enable the development of interpretive frameworks regarding the clinical actions that should follow as measurement thresholds are crossed, which is an essential step in achieving broad clinical adoption of novel digital measures. The substantial financial benefits that might be achieved when such precompetitive collaborations lower costs are suggested by our results when mean implementation cost is replaced in the model by the much lower median cost.

The nature of digital measures and digital measurement tools may also influence the total costs and returns associated with investments. Organizations that choose to invest in developing new sensor products will accrue much greater costs than organizations who partner with hardware product developers and measure developers. In many cases, developers of digital measures and digital measurement products already have evidence of verification, analytical validation, and/or clinical validation, so those aspects of measure development may not need to be performed for subsequent use [19].

## LIMITATIONS

This study was limited by some relatively small sample sizes. The implementation cost analysis in particular was constrained by the proprietary nature of the data; many individuals invited to complete the survey (across both trial sponsors and digital measure developers) responded that they were unable to share the requested data as it was deemed proprietary and could not be disclosed. This limitation is a consequence of prevailing industry practices that must change to establish industry benchmarks that characterize so many other facets of clinical research.

The full economic analyses in this study were necessarily restricted to three therapeutic areas due to limited data available for other therapeutic areas. As the application of digital endpoints expands over time, as we expect, one will be able to assess the value of using digital endpoints more broadly. Such an expansion of the analysis requires that data on digital endpoint use continue to be collected in a repository such as the Library of Digital Endpoints. It could also be helpful if clinical trial registries such as ClinicalTrials.gov begin collecting information on digital endpoints. Beyond indicating if digital endpoints have been employed, this could also be a useful vehicle for expanding our understanding of the impact of adopting digital endpoint strategies if the details of such use are reflected in study results when they are made available, as has been suggested elsewhere [20].

It may reasonably be argued that the impacts of utilizing digital endpoints in clinical trials on trial sizes and trial durations are driven by those digital endpoints that are positioned as primary outcomes. The sample sizes for digital endpoints here are too small to analyze trials with primary outcome digital endpoints by phase and therapeutic area. However, if trials where no digital endpoints are positioned as a primary outcome have little to no effect on trial sizes and durations, their inclusion in the analyses of digital endpoint trials argues that our net benefit results are conservative. The data we have suggest that this is the case. When aggregating across all phases and therapeutic areas the mean trial sizes and durations for both therapies and devices are lower for digital endpoint trials where at least one of the digital endpoints is a primary outcome compared to digital endpoint trials where none of the digital endpoints are positioned as primary outcomes.

Given the availability of data, we were limited to examining just two metrics that can be a source of value (trial duration and trial enrollment). Future research must also identify the additional costs and benefits to evaluate the total absolute value that developing and deploying digital endpoints brings to the drug development industry (e.g., participant, investigator, and site burden; accessibility of clinical trials; downstream reimbursement decision making; internal decision making and other potential uses). Furthermore, to drive the maturity of digital measurement capabilities across the field, sustainable methods for monitoring and benchmarking the continued growth of digital endpoints as a new tool in the toolbox for clinical development programs must be established. Additional frameworks may also support organizations in evaluation of eNPV and ROI on a case-by-case basis.

## CONCLUSION

This study demonstrates that, despite the need for substantial upfront investment, digital endpoints are yielding substantial eNPV and ROI to the drug development industry. For phase 2 base case analyses, the increase in eNPV from employing digital endpoints varied from $2.2 million to $3.3 million, with ROIs between 32% to 48%, per indication for the three therapeutic areas analyzed. Positive financial impacts were substantially higher for phase 3 trials. In phase 3 base case analyses, the increase in eNPV ranged from $27 million to $48 million, with returns that were four to seven times the investment. Digital endpoints do not simply promise to address some of the greatest challenges facing today’s clinical trials enterprise but are actively capturing this value today.

These findings serve as an important reminder that scientific evaluation of innovative approaches in drug development must be accompanied by economic analyses to establish that those innovations that provide value to sponsor organizations continue to receive investment to capture these benefits for the industry and the patients who depend upon them. Findings such as these can drive acceptance of digital endpoints in the decision-making process of relatively risk-averse organizations. Widespread adoption of digital endpoint strategies can not only transform drug development, but also yield substantial social benefits, as safe and effective treatments will reach patients sooner, and, if digital measures are also adopted widely in clinical practice, patient adherence and clinical outcomes can improve.

## Author contributions

Substantial contributions to the conception or design of the work (JD, ZS, AD, JG, SV); substantial contributions to the acquisition, analysis, or interpretation of the work (JD, ZS, AD, BH); drafting the work (JD, SV, JG, BH, TM, UG); revising the work critically for important intellectual content (JD, ZS, AD, SV, JG, TM, UG, LL, UC, DK, LM, KG, BH); final approval of the version to be published (JD, ZS, AD, SV, JG, TM, UG, LL, UC, DK, LM, KG, BH); agreement to be accountable for all aspects of the work in ensuring that questions related to the accuracy or integrity of any part of the work are appropriately investigated and resolved (JD, ZS, AD, SV, JG, TM, UG, LL, UC, DK, LM, KG, BH).

## Data Availability

All data produced are available online at:
https://aact.ctti-clinicaltrials.org/
https://dimesociety.org/get-involved/library-of-digital-endpoints/

## Footnotes

1 Device clinical development typically does not follow the sequential phase framework that marks drug development. Thus, for device trials we do not show comparative results by clinical phase.

2 Descriptive statistics for the number of countries in which the trial is conducted differed little between trials where digital endpoints were employed and where they were not (Supplemental Figures S2-S7).

