## Supplemental data file for "ASSESSING THE NET FINANCIAL BENEFITS OF EMPLOYING DIGITAL ENDPOINTS IN CLINICAL TRIALS"

#### Figure S1. Therapeutic Class Distribution for Trials with Digital Endpoints

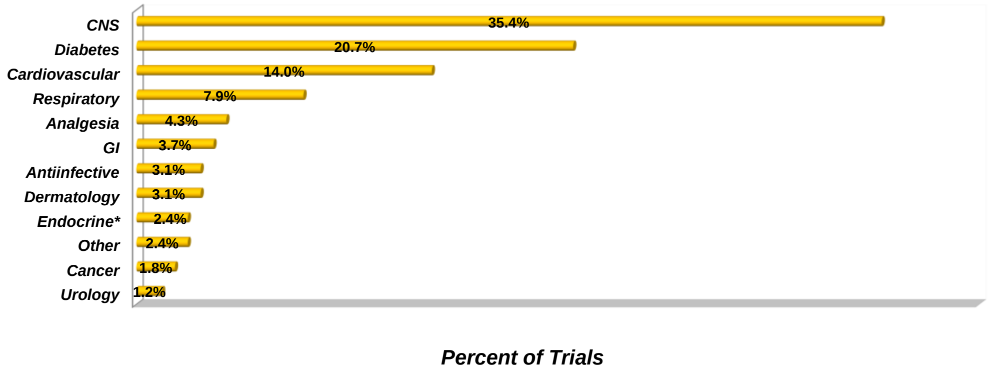


*** Excludes diabetes trials**

**n = 164 trials**

#### Figure S2. Distribution of Number of Countries per Trial in ClinicalTrials.gov and DiMe Digital Endpoints Data

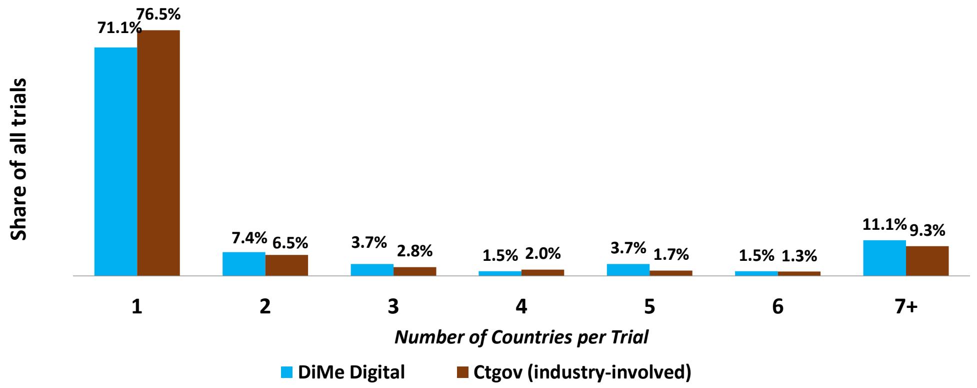


DiMe digital: n = 135 trials
ClinicalTrials.gov (industry-involved): n = 126,339 trials

#### Figure S3. Mean Number of Countries per Trial in ClinicalTrials.gov and DiMe Digital Endpoints Data

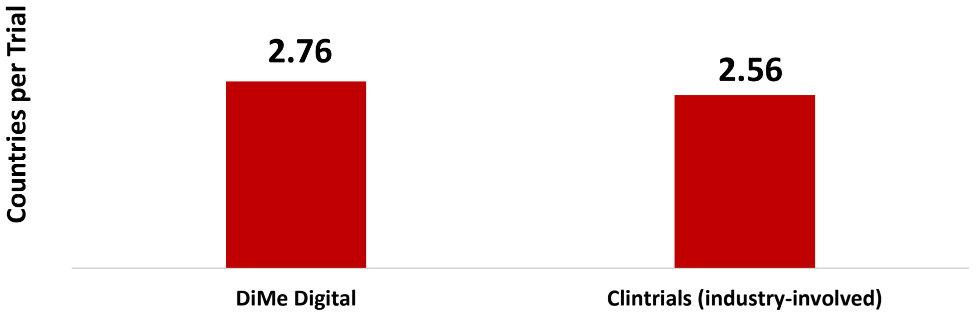


DiMe digital: n = 135 trials
ClinicalTrials.gov (industry-involved): n = 126,339 trials

### Figure S4. Single Country Share of Trials in ClinicalTrials.gov by Trial Phase

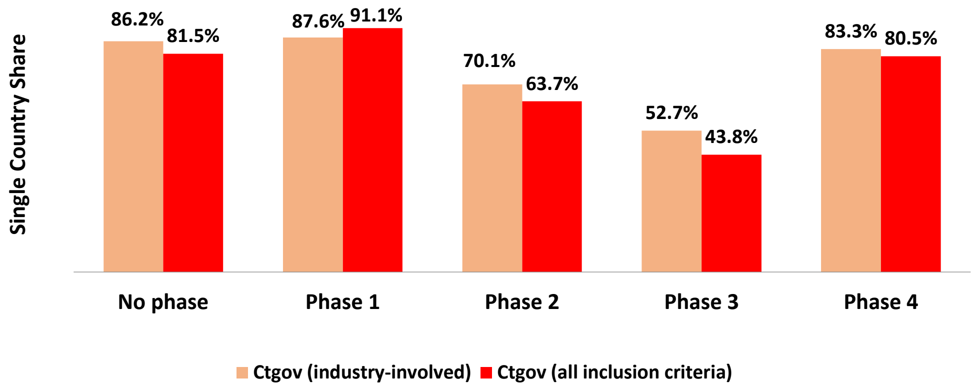


ClinicalTrials.gov (industry-involved): n = 126,339 trials

ClinicalTrials.gov (all inclusion criteria): n = 10,394 trials

#### Figure S5. Mean Number of Countries per Trial in ClinicalTrials.gov

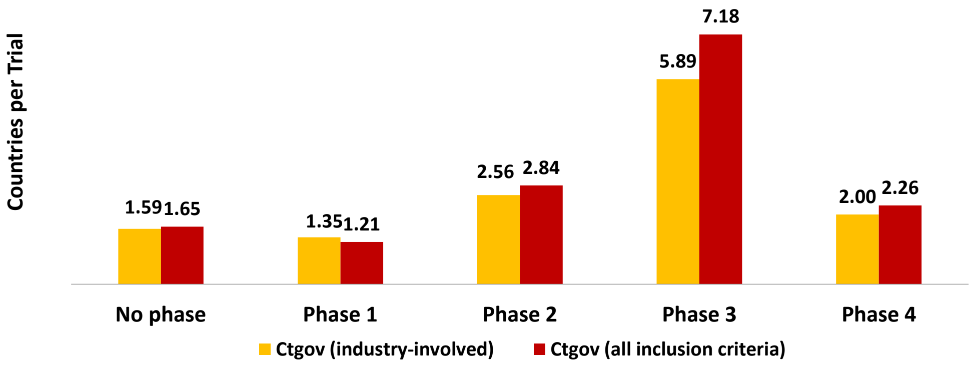


Clinicaltrials.gov (industry-involved): n = 126,339 trials
Clinicaltrials.gov (all inclusion criteria): n = 10,394 trials

#### Figure S6. Geographic Distribution of Trials in DiMe Digital Endpoints and ClinicalTrials.gov Data by Region

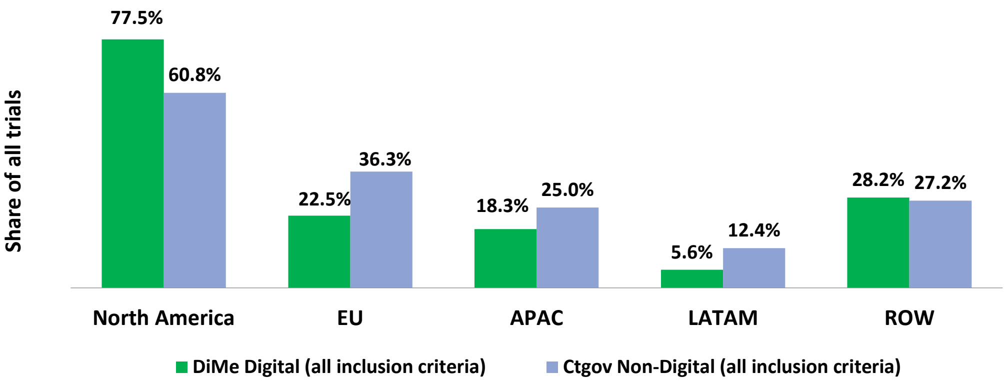


DiMe digital (all inclusion criteria): n = 71 trials

ClinicalTrials.gov (all inclusion criteria): n = 10,399 trials

Percentages add to more than 100 because some trials are multi-regional

#### Figure S7. Distribution of Number of Regions per Trial for DiMe Digital Endpoints and ClinicalTrials.gov Data

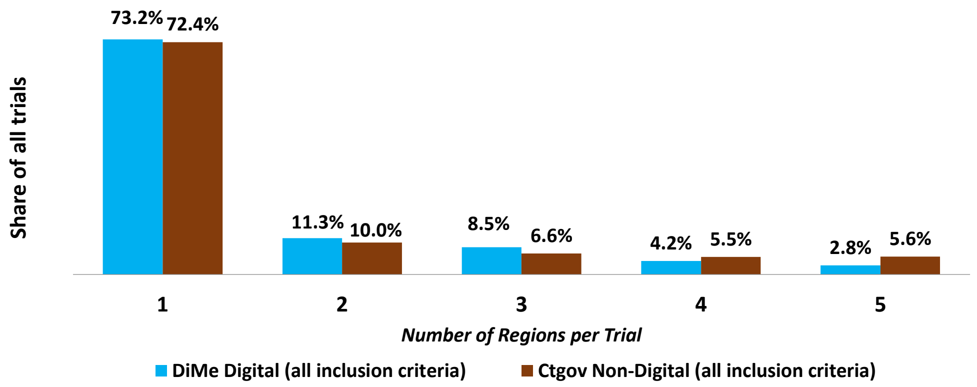


DiMe digital (all inclusion criteria): n = 71 trials
ClinicalTrials.gov (all inclusion criteria): n = 10,399 trials

#### Figure S8. Distribution of Digital Endpoints by Trial Outcome Positioning

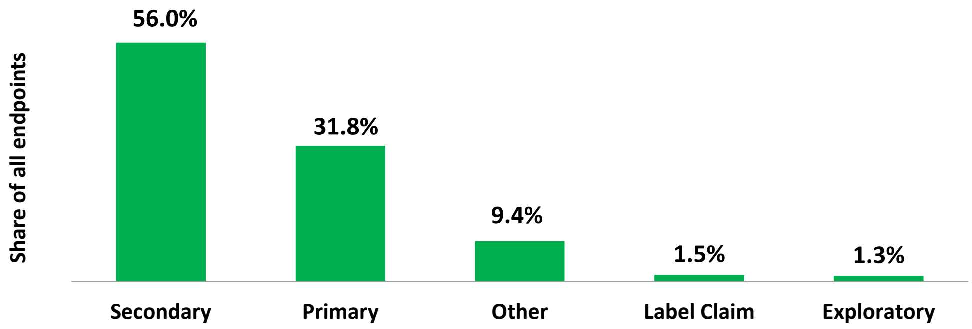


n = 363 digital endpoints

### Figure S9. Distribution of Technology Types Used to Measure Digital Endpoints

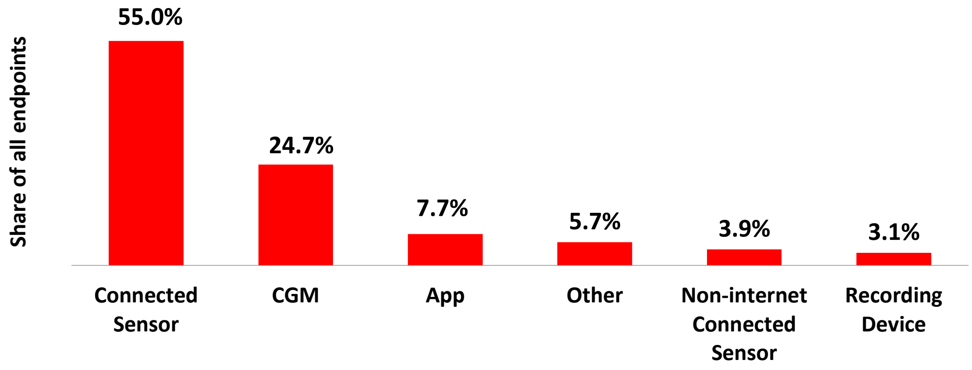


n = 389 digital endpoints

#### Figure S10. Number of Digital Endpoints per Trial

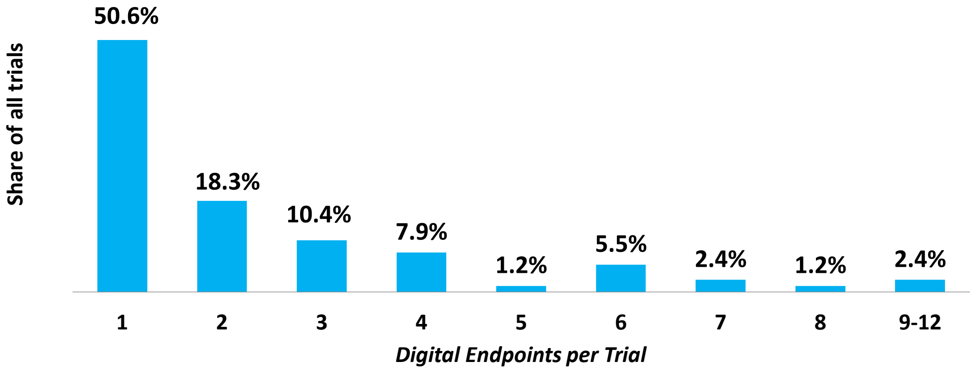


n = 164 trials

#### Figure S11. Number of Sites per Trial in ClinicalTrials.gov and DiMe Digital Endpoints Data

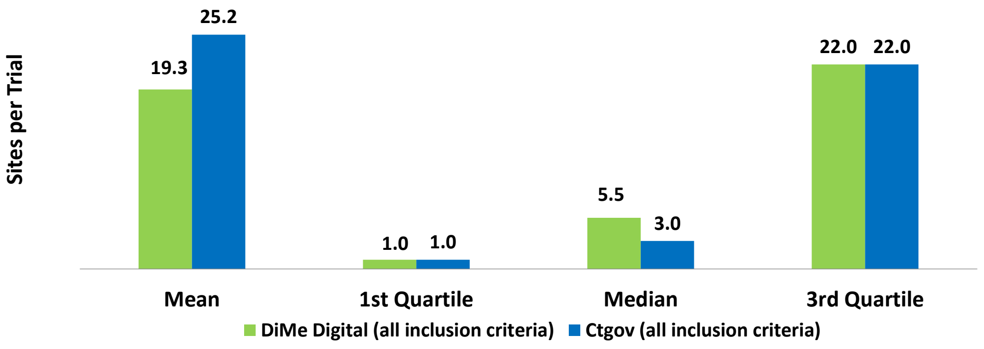


DiMe digital: n = 72 trials
ClinicalTrials.gov: n = 10,408 trials

#### Figure S12. Number of Sites per Trial in ClinicalTrials.gov and DiMe Digital Endpoints Data (extreme outliers excluded – IQR rule)

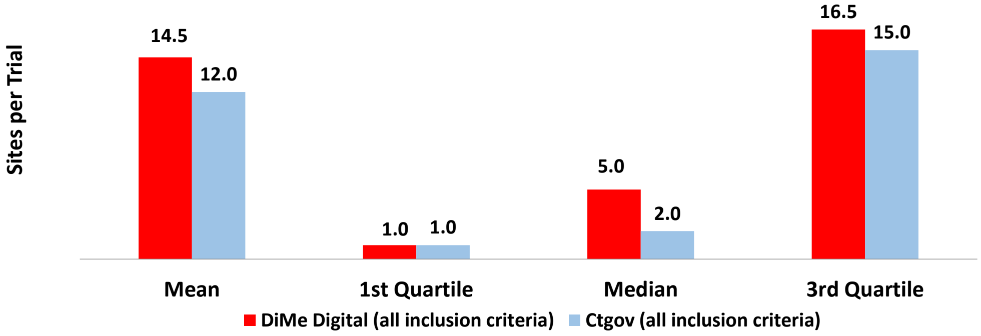


DiMe digital: n = 68 trials
ClinicalTrials.gov: n = 9,602 trials

#### Figure S13. Total Number of Endpoints per Trial in ClinicalTrials.gov and DiMe Digital Endpoints Data

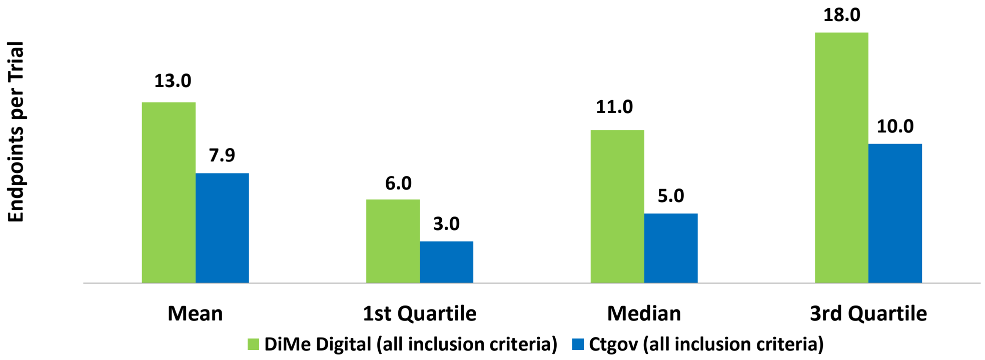


DiMe digital: n = 79 trials
ClinicalTrials.gov: n = 11,363 trials

#### Figure S14. Total Number of Endpoints per Trial in ClinicalTrials.gov and DiMe Digital Endpoints Data (extreme outliers excluded – IQR rule)

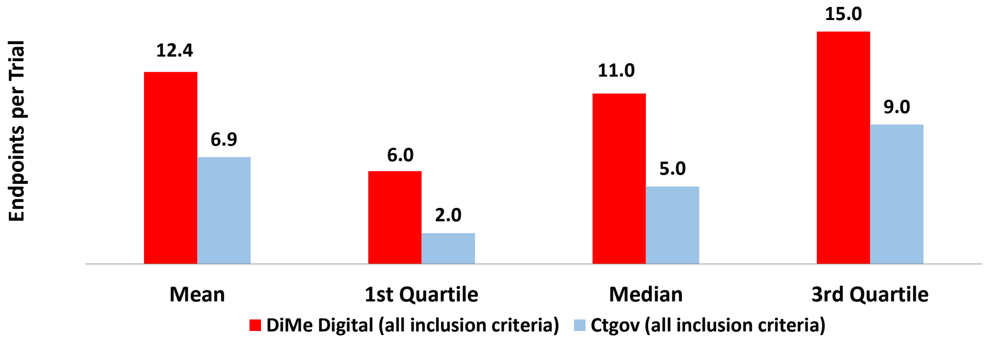


DiMe digital: n = 78 trials
ClinicalTrials.gov: n = 11,104 trials

Table S1. Final Model Regression Coefficients for Trial Time and Size by Phase

| | Dependent Variable | | | |
|---|---|---|---|---|
| | Log of phase 2 duration | Log of phase 3 duration | Phase 2 enrollment | Log of phase 3 enrollment |
| **Explanatory variables** | | | | |
| Intercept | 2.2402 | 2.53354 | 167.12977 | 5.48882 |
| Digital | -0.27431 | -0.24874 | -15.23932 | -0.12445 |
| CNS | 0.24135 | 0.27220 | -21.5209 | -0.12081 |
| Cardio | 0.07549 | 0.18121 | -37.85677 | -0.54550 |
| Year | 0.02228 | 0.01915 | -2.98029 | 0.01075 |
| Sites | 0.00377 | 0.00135 | --- | --- |
| Endpoints | --- | 0.00281 | 0.54203 | 0.01196 |
| | N=1,192; F=16.2; p<0.0001 | N=1,303; F=31.5; p<0.0001 | N=1,244; F=9.7; p<0.0001 | N=1,378; F=14.1; p<0.0001 |

**Table S2. Mean Values for Continuous Independent Variables for Cycle Time and Trial Size Regressions for Phase 2 and Phase 3**

| | ***Cycle Time*** | | |
|---|---|---|---|
| | Number of sites | Number of endpoints | Year* |
| Phase 2 | 18.5 | 9.0 | 13.8 |
| Phase 3 | 73.9 | 10.4 | 12.6 |
| | ***Trial Size*** | | |
| | Number of Sites | Number of Endpoints | Year* |
| Phase 2 | 16.8 | 8.9 | 13.6 |
| Phase 3 | 57.0 | 10.2 | 12.2 |

* 2005 = year 0

**Figure S15. Sponsor Costs from Implementing, Developing, and Validating Digital Endpoints in Clinical Trials (2023 $)**

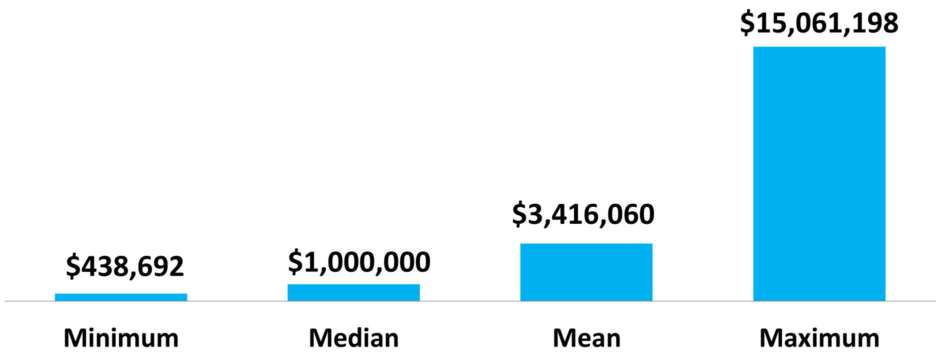


n = 11

### Table S3. Key Parameters and Data Sources

| Parameter | Data Source | Parameter | Data Source |
|---|---|---|---|
| Development and review times | DiMasi et al., *J Health Econ* 2016;47:20-33 and CSDD protocol database | Peak sales and years to peak | Cortellis pipeline database (consensus analyst forecasts) |
| Development costs | DiMasi et al., *J Health Econ* 2016;47:20-33 and CSDD protocol database | Effective tax rate | Public financial data for top 10 pharma firms |
| Phase success rates | BIO/Informa/QLS, Feb 2021 | Digital endpoint implementation cost | CSDD/DiMe Sponsor and Developer Cost Survey |
| Cost of capital | DiMasi et al., *J Health Econ* 2016;47:20-33 | Change in trial duration | ClinicalTrials.gov and DiMe databases |
| Approved supplemental indications | Drugs@FDA | Change in trial size | ClinicalTrials.gov and DiMe databases |

#### Figure S16. Mean Number of FDA-approved Indications by Therapeutic Area and Approval Period

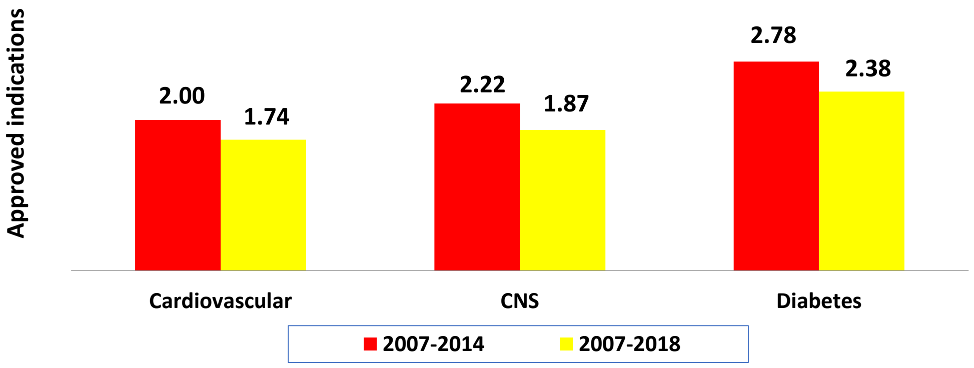

**Table S4. Development Risk Parameter Values for eNPV Analysis (indication transition probabilities)**

| Transition | Cardiovascular | Endocrine | Neurology |
|---|---|---|---|
| Phase 2 to Phase 3 | 21.0% | 26.6% | 26.8% |
| Phase 3 to Regulatory Review | 55.2% | 66.2% | 53.1% |
| Regulatory Review to Approval | 82.5% | 86.3% | 86.7% |
| Phase 2 to Approval | 9.6% | 15.2% | 12.3% |
| Phase 3 to Approval | 45.5% | 57.1% | 46.0% |

Source: Clinical Development Success Rates and Contributing Factors, 2011-2020, Biotechnology Innovation Organization (BIO), Informa Pharma Intelligence, QLS Advisors, url: https://www.bio.org/clinical-development-success-rates-and-contributing-factors-2011-2020

**Table S5. Relative* Clinical Phase to Phase Durations and Phase R&D Costs**

| | Cardiovascular | Diabetes | CNS |
|---|---|---|---|
| Phase 2 to Phase 3 relative duration | 90.4% | 98.9% | 102.0% |
| Phase 3 to regulatory review relative duration | 110.5% | 76.1% | 95.0% |
| Phase 2 relative cost | 94.9% | 97.1% | 163.1% |
| Phase 3 relative cost | 46.8% | 62.9% | 119.7% |

* Relative to overall averages for drug in general

Source for overall averages and relative durations and costs: DiMasi et al., *Journal of Health Economics* 2016;47:20-33 and Tufts CSDD Protocol Complexity Benchmark Database

### Figure S17. Sensitivity Analyses for Phase Duration and Trial Size by Phase and Therapeutic Area (at median implementation cost)

Figure S17a. Phase 2 digital endpoint ROI by reduction in trial duration (at base case decrease in trial size)

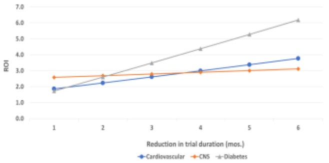


Figure S17b. Phase 2 digital endpoint ROI by percentage reduction in trial size (at base case decrease in trial duration)

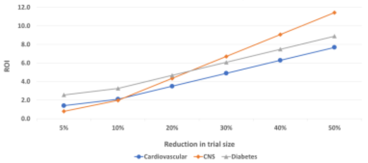


Figure S17c. Phase 3 digital endpoint ROI by reduction in trial duration (at base case decrease in trial size)

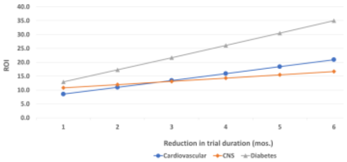


Figure S17d. Phase 3 digital endpoint ROI by percentage reduction in trial size (at base case decrease in trial duration)

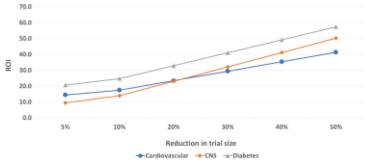
